# The impact of strict public health measures on COVID-19 transmission in developing countries: The case of Kuwait

**DOI:** 10.1101/2020.05.03.20089771

**Authors:** Abdullah A. Al-Shammari, Hamad Ali, Barrak Alahmad, Faisal H. Al-Refaei, Salman Al-Sabah, Mohammad H. Jamal, Abdullah Alshukry, Qais Al-Duwairi, Fahd Al-Mulla

## Abstract

**Background:** Many countries have succeeded in curbing the outbreak of COVID-19 by employing strict public health control measures. However, little is known about the effectiveness of such control measures in curbing the outbreak in developing countries. In this study, we seek to assess the impact of various outbreak control measures in Kuwait to gain more insight into the outbreak progression and the associated healthcare burden.

**Methods:** We use a SEIR mathematical model to simulate the epidemic outbreak of COVID-19 in Kuwait with additional testing and hospitalization compartments. We use a NBD observational framework for confirmed case and death counts. We forecast model trajectories and assess the effectiveness of public health interventions by using maximum likelihood to estimate both the basic and effective reproduction numbers.

**Results:** Our results indicate that the early strict control measures had the effect of delaying the intensity of the outbreak but were unsuccessful in reducing the effective reproduction number below 1. Forecasted model trajectories suggest a need to expand the healthcare system capacity to cope with the associated epidemic burden of such ineffectiveness.

**Conclusion:** Strict public health interventions may not always lead to the same desired outcomes, particularly when population and demographic factors are not accounted for as in the case in some developing countries. Real-time dynamic modeling can provide an early assessment of the impact of such control measures as well as a forecasting tool to support outbreak surveillance and the associated healthcare expansion planning.

**SUMMARY BOX:** *What is already known on the subject?:* Evidence is accumulating about the positive impact of various strict public health interventions on the transmission of COVID-19 in the developed world. Currently, however, many developing countries are still struggling to control and suppress the initial wave of the outbreak. In particular, less attention is given to assessing the impact of taking similar strict control measures.

*What does this study add?:* Our modeling study provides the first evidence showing how the imposition of strict public health measures has not led to a reduction in COVID-19 transmission in Kuwait. It highlights the importance of performing systematic epidemiological and public health investigations of the population factors which may limit the effectiveness of standard public health interventions in developing countries. It also emphasizes the utility of adopting dynamic modeling approaches for intervention assessment and healthcare capacity re-adjustment at the earliest stages of the outbreak.

## INTRODCUTION

In early December 2019, a cluster of pneumonia cases of unknown cause were reported in Wuhan, China [1]. Later the pathogen was identified and named, Severe Acute Respiratory Syndrome Corona Virus 2 (SARS-CoV2), an enveloped single strand RNA β-coronavirus with a genome of almost 30 thousand bases [2]. Since then the virus has been spreading rapidly all over the globe with the World Health Organization (WHO) confirming Corona Virus Disease 2019 (COVID-19) as Pandemic on March 11^th^, 2020. With the number of cases reaching a staggering 3.5 million cases in more than 200 countries and a death toll exceeding 250 thousand [3], coordinated worldwide efforts are needed to prepare healthcare systems to cope with this unprecedented challenge. While some countries showed a degree of resilience and capacity to deal with the progression of COVID-19, in others the burden on healthcare systems was overwhelming leading to catastrophic consequences.

Many developing countries are struggling to control the COVID-19 outbreak, since little is known about the effectiveness of public health measures taken by countries with smaller populations and unique demographic profiles. Kuwait is a small wealthy country with a population of nearly 4.7 million people. On February 24^th^, Kuwait recorded the first confirmed cases of COVID-19 in four passengers arriving from Iran. Since then, the Ministry of Health have confirmed more than 4000 COVID-19 cases and 26 deaths. Kuwait has implemented early strict control measures in attempt to contain the spread of SARS-CoV2 including: closure of schools, universities, governmental offices and non-essential businesses; full border lockdown, partial curfew and geographic isolation of areas experiencing wide community transmission (Figure 1). The situation in Kuwait was further complicated by a remarkable repatriation operation to bring back more than 50,000 Kuwaiti citizens from around the world by May 7th, 2020. The government is implementing home and institutional quarantine measures to limit virus transmission from arrivals. Despite these early and stringent control measures, community transmission remains observed as manifested by the apparent acceleration of case and death numbers well beyond the anticipated period of slowdown. Hence it is unclear how the outbreak will unfold in the next few months as recent contact-tracing measures highlighted the widening community transmission. In addition, the polymerase chain reaction (PCR) testing seems to be constrained by a global shortage of testing kits and reagents posing a threat to smaller countries to get necessary testing tools who now found themselves competing internationally with other countries. In anticipation of the unfolding of such circumstance it becomes necessary to forecast the potential burden it may incur on local healthcare systems.

**Figure 1.**
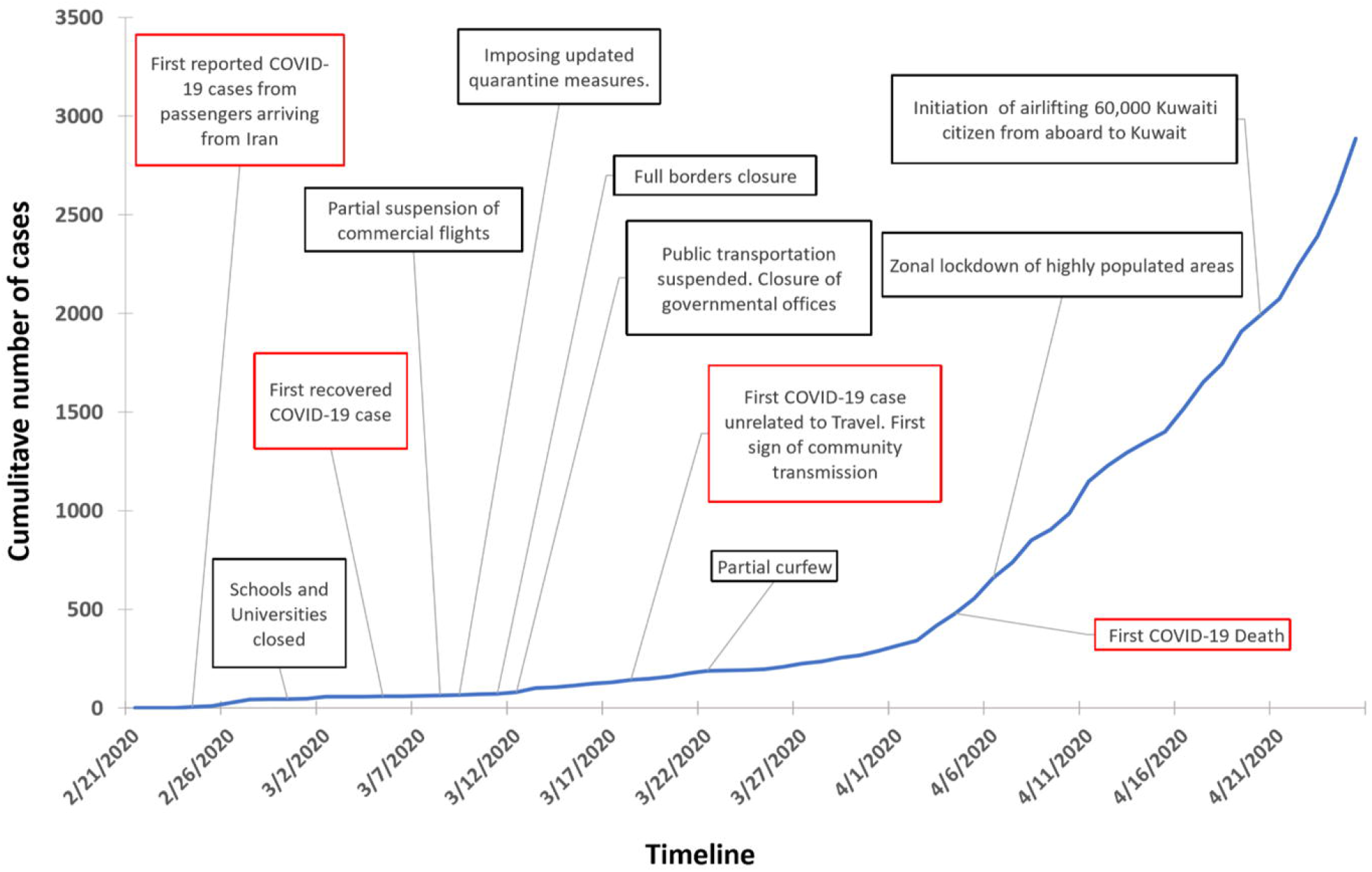
Cumulative number of reported COVID-19 cases in Kuwait along with a timeline of events.

Forecasting the outbreak dynamics of COVID-19 cases in Kuwait is crucial to estimating, well in advance, the potential burden on the healthcare system. These epidemic outbreak dynamics are typically investigated by employing mathematical models of infectious disease transmission dynamics such as the classic Susceptible-Infective-Recovered (SIR) or Susceptible-Exposed-Infective-Recovered (SEIR) epidemiological models [4-6]. Such models play a pivotal role in understanding the epidemic characteristics of an infectious disease outbreak [7], as well as in assessing the impact of various interventions on the spread of the disease [6, 8-11]. Indeed, various low income countries are previously used these models to help inform their epidemic containment policies [6]. Different generalized versions of these mathematical models can provide more detailed mechanisms for the epidemic dynamics (e.g. mode of transmission, quarantine dynamics, testing scope, and hospitalization dynamics). A widely adopted model for characterizing the epidemic outbreak SARS-CoV-2 is the SEIR model [5]. However, under-reporting in daily case numbers due to nonoptimal testing poses a significant challenge to understanding the trends associated with COVID-19 progression by public health authorities [12]. One way to mitigate the impact of this structural limitation is by fitting a dynamic transmission model to daily numbers of incident cases of infections as well as reported deaths [13].

In this study, we model the progression of the COVID-19 outbreak in Kuwait by developing a generalization of the SEIR model that is informed by two local mechanisms; a delay period during which suspected COVID-19 individuals are tested, identified and hospitalized, and different severity of illness (ranging from recovered asymptomatic to needing critical care). We then calibrate the model by applying a maximum likelihood framework using incident cases of infections and reported deaths [14]. We use this framework to assess the effectiveness of public health interventions and forecast the associated healthcare burden in Kuwait.

## METHODS

### Mathematical model

We use a deterministic compartmental model for infectious disease transmission with additional compartments to describe the dynamic burden on the healthcare system (Figure 2). Our model simulates SEIR, testing and hospitalization dynamics and can be described by the following set of differential equations:

**Figure 2.**
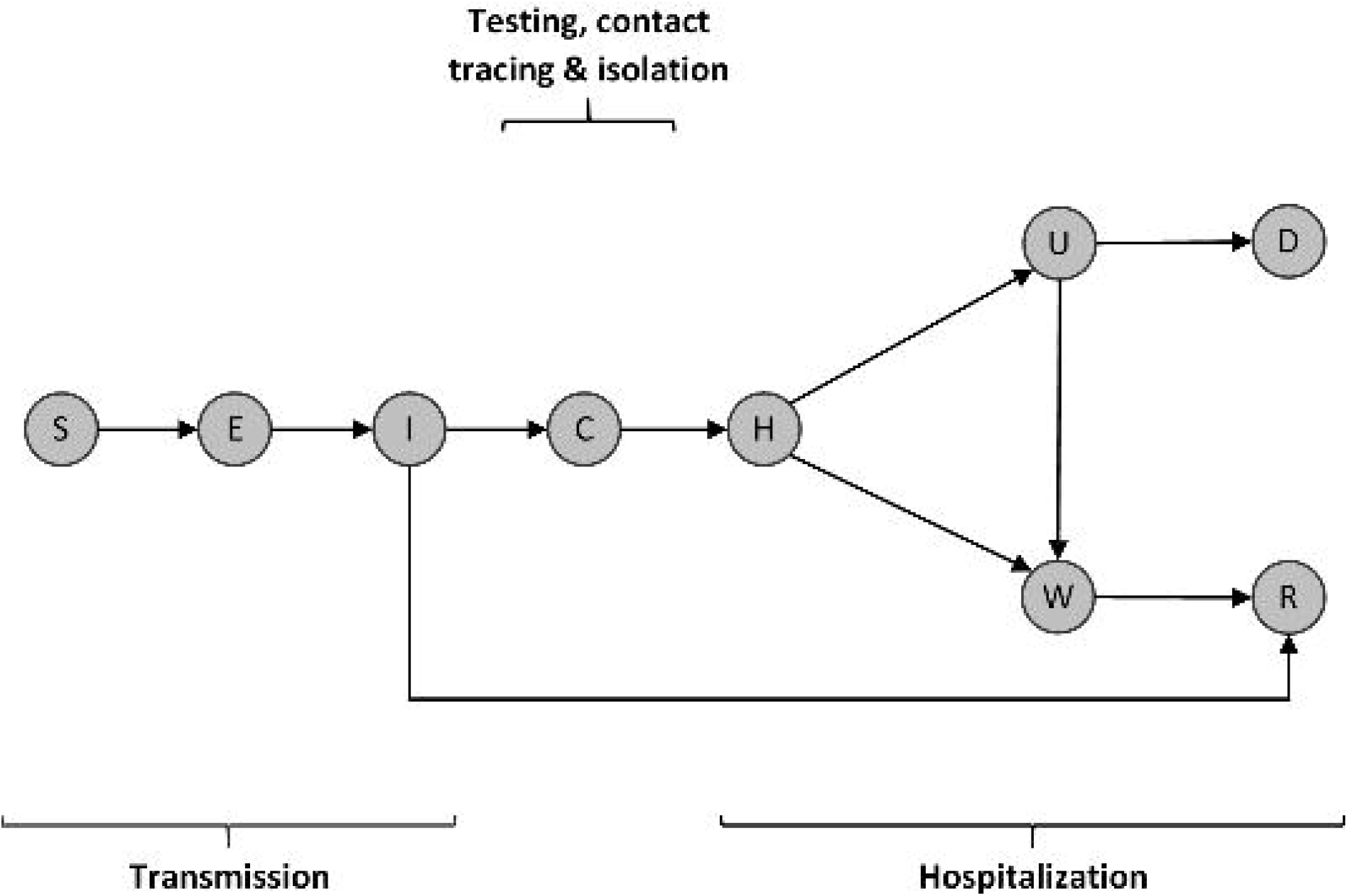
Schematic diagram of the CoVID-19 transmission model. Individuals (S) susceptible to the virus become infected by infectious individuals (I). They then move through a latent period (E) before becoming infectious (I). Infectious individuals can either move through a detection period (C) or eventually recover without symptoms. Confirmed infectious individuals move through an initial hospitalization period (H) after which they are admitted to either an isolation ward (W) or an intensive care unit (U). Intensive care patients may recover and be sent to an isolation ward W or ultimately die (D). Isolated patients move through a recovery period (R), where they are assumed to be immune to the disease, at least in the medium term.

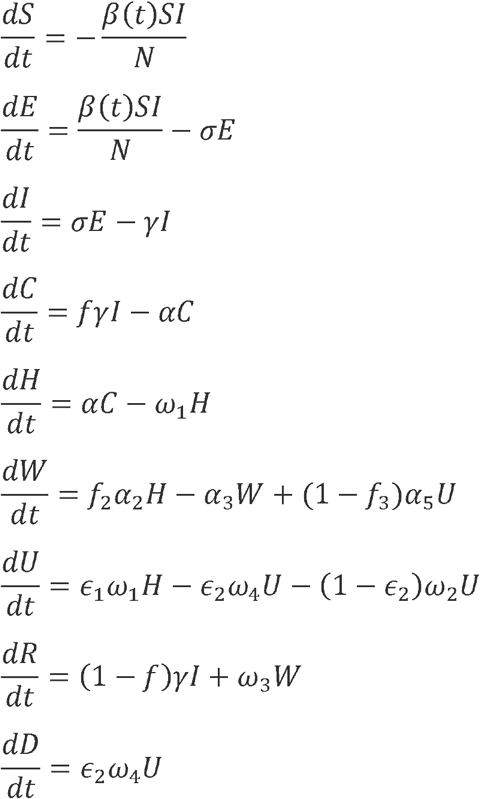

Upon infection, individuals that are susceptible to the virus (S) become exposed but non-infectious carriers (E) and later infectious (I). A fraction of infectious individuals may remain undetected and ultimately enter into a recovered class (R), while the remaining fraction end up being detected (C) by some form of clinical testing or diagnosis. Detected individuals are sent to hospitals (H) where they are admitted to either an isolation ward (W) or an intensive care unit (U) based on the intensity of their symptoms. Intensive care patients either die (D) or get sent to an isolation ward to stay until full recovery (R) (Figure 2).

The progression through the different compartments in our model is characterized by key time periods which describe the dynamic transmission of infection, case detection, patient care and hospitalization, and recovery or death: the average durations of viral latency (1/σ), carrier infectiousness (1/γ), onset-to-hospitalization (1/α), onset-to-death, hospitalization-to-discharge, and ICU-stay. The fraction of individuals who end up being detected (*f*) is related to the case fatality rate. Since the transmission rate β is affected by the implementation of control measures, we take it as a function of time *β* (*t*) = *β*_0_ *k* (*t*), where *β*_0_ is the transmission rate without control measures (baseline) and *k*(*t*) is a positive scaling factor by which interventions may reduce the transmission rate. In other words, *k* values smaller than 1 characterize a more effective intervention in curbing the epidemic. Here *k* = 1 indicates an ineffective or absent control measure, with values greater implying improper control measures.

We remark here that the modelling was made possible despite the lack of population health data such as country demographics and associated person-person contact structure. We assumed homogeneous mixing not accounting for age-structure nor the risk associated with comorbidities. Nonetheless, these can be easily incorporated into our model once detailed data become available.

### Data & parameters

Numbers of confirmed infection and death cases were collected from daily reports from the European Center for Disease Control [15]. All our data sources are outlined in table 1. The key time durations in our model were fixed to values obtained from published datasets as well as unpublished local hospitalization data. The mean durations of latency, incubation and infectiousness of SARS-CoV-2 ware based on the reported cases from the COVID-19 outbreak in Singapore and Tianjin, China [16].

**Table 1.** Model parameters. Rate of testing and the proportion of cases being tested remain largely unknown at this stage but are expected to increase over time as health authorities increase their laboratory testing capacity. The hospital care data were provided by colleagues from the Ministry of Health, Kuwait.

| Symbol | Definition | Default Values | Justification |
| --- | --- | --- | --- |
| $N$ | Total population of Kuwait | 4,776,000 | PACI, Kuwait |
| $S_0$ | Susceptible subpopulation | 500,000 | MOH, Kuwait |
| $R_0$ | Basic reproduction number | 1.5 – 3.5 | [21] |
| $\kappa$ | Factor for transmission reduction | 0.5 | |
| $\sigma^{-1}$ | Latent period | 2 days | [16] |
| $\gamma^{-1}$ | Infectious period | 3.2 days | [16] |
| $\alpha^{-1}$ | Onset-to-hospitalization period | 2 days | Unpublished data |
| $\omega_1^{-1}$ | Initial hospitalization period | 6 days | Unpublished data |
| $\omega_2^{-1}$ | Mean ICU duration until recovery | 8.5 days | Unpublished data |
| $\omega_3^{-1}$ | Mean isolation ward duration | 10 days | Unpublished data |
| $\omega_4^{-1}$ | Mean ICU duration until death | 10.5 days | Unpublished data |
| $f$ | Proportion of tested & reported daily cases | 0.12 | [22] |
| $\varepsilon_1$ | Proportion of patients admitted to ICU | 0.075 | Unpublished data |
| $\varepsilon_2$ | Proportion of ICU patients with death outcome | 0.25 | Unpublished data |
| CFR | Case fatality ratio | 1.4% | [12] |

We assume that a single case started the outbreak on February 24^th^, 2020, which coincides with the date of the first reported case of COVID-19 in Kuwait. The total population of Kuwait (N) is about 4,776,000. We note here that our model parameter estimation is insensitive to the number of susceptible individuals as long as the number of cases is small relative to N. We take the initial unprotected susceptible population to be 500,000 which is the effective number of individuals who account for the majority of local community transmission in Kuwait. This estimate is consistent with the assumptions that 1) the majority of the population has been protected by the stay-at-home orders, 2) most community transmission cases are localized to certain geographic areas, and 3) children younger than 18 years old represent a very small percentage of the total number of infected patients. However, we also model 1,500,000 susceptible individuals and show the corresponding results.

To assess the impact of control interventions, we assume *k* (*t*) = 1 prior to the implementation of a partial lockdown on March 22, 2020 (Figure 1). We then estimate *K* and the baseline transmission rate *β*_0_ by employing a maximum likelihood framework [14]. To derive the maximum likelihood estimates (MLE) of our unknown parameters we assume the daily numbers of incident infections are detected according to a negative binomial distribution (NBD). We additionally assume the daily number of incident deaths are drawn from a similar distribution. Then optimization was carried out using the Nelder-Mead method [17] on the combined minus log-likelihood function.

The uncertainty of parameters was represented by quantile-based credible intervals (CI). We use the asymptotic normality of MLE to account for such uncertainty through deriving simulation-based 95% CIs for the model curves [18]. Simulations were run 10,000 times based on random draws of the unknown model parameters from a normal distribution 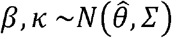. Here 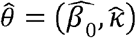 are maximum likelihood estimates and *Σ* is the variance-covariance matrix associated with them. Given the parameterization of our transmission model, these parameters permit a model-based estimation of the basic and effective reproduction numbers. In particular, in each simulation run an R_0_ value is drawn from a range of values (Table 1) as an initial point to kick start the parameter search algorithm. Our transmission model was fitted to estimate key transmission parameters. The maximum likelihood estimates of the baseline and effective transmission rates, *β*_0_ and *β*_e_ were used compute the basic and effective reproduction numbers via these formulas

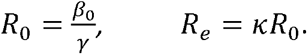

All simulations, parameter estimation and model calibration were run in the R software [19].

## RESULTS

Our estimated basic reproduction number is *R*_0_ = 1.43 (95% CI: 1.33–1.58). Interestingly, the MLE of the factor by which control measures reduce transmission was estimated at *k*= 1 (95% CI: 0.9998–1). This corresponds to an effective reproduction number *R*_e_ = *R*_0_, which is consistent with reports from the Centre for Mathematical Modelling of Infectious Diseases [20].

We remark here that our model-based estimates of the reproduction numbers, which directly influence the prevalence of the epidemic, depend on the values we adopt for the incubation and infectious periods (Table 1). In particular, larger periods are expected to lead to higher values for the reproduction numbers. For example, we find *R*_t_ = *R*_0_ = 1.97 (95% CI: 1.85– 2.12) if we change the incubation and infectious periods to 5 and 6 days, respectively.

Our parameter estimations captured the variation around the observed number of reported cases and deaths to project a posterior distribution of the expected numbers. Our projected trajectories and their 95% credible intervals were able to capture the early slow increase in observed cases, hospitalization and deaths (Figure 3).

**Figure 3.**
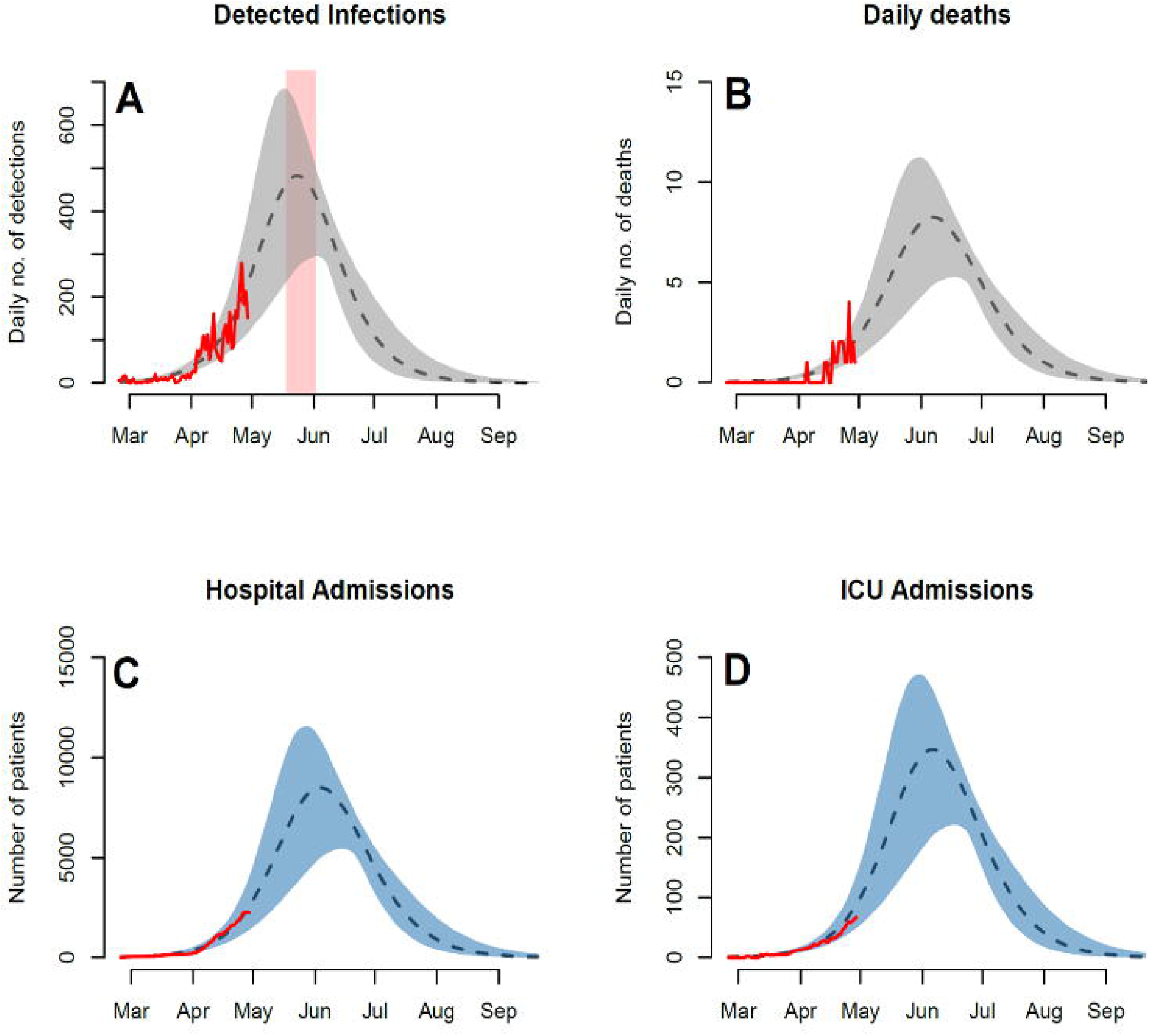
Observed and forecasted trajectories assuming 500,000 unprotected susceptible individuals. Observed and projected daily numbers of A) incident infections, B) death cases, C) general hospital admissions and D) ICU admissions. Red rectangular ribbon highlights the projected time-window of the epidemic peak. Red lines represent the reported data. Black dashed lines represent model projections based on MLE of unknown parameters with shaded ribbons representing 95% credible interval on new observations. We note here that the observed cases and their projections only represent a fraction of the actual and model prevalence. This is based on our assumption of under-reporting and the presence of asymptomatic individuals in the population.

Under the current testing rate, capacity and scope, the model projects the daily numbers of reported cases to peak around 480 (95% CI: 300—680) by the second half of the month of May. In terms of the burden on the healthcare system, our model projects peak hospital admission of 8000 patients (95% CI: 5000—12000) with ICUs projected to peak around 350 patients (95% CI: 220—480). At these rates the model projects a peak daily mortality around 8 deaths (95% CI: 5—12). We additionally explored a scenario that simulates an expansion in the size of the susceptible subpopulation by a factor of three (Figure 4). A summary of the projected epidemic and healthcare burdens is presented in Table 2.

**Table 2.** Projected epidemic and healthcare burdens. Burden projections based on model simulations are presented. Uncertainty is represented by with 95% credible intervals.

| Expected Burden | 500,000 Susceptibles | 1,500,000 Susceptibles |
| --- | --- | --- |
| Max reported cases | 480 (300–680) | 1400 (800–2000) |
| Max hospital occupancy | 8000 (5000–12000) | 25000 (15000–35000) |
| Max ICU occupancy | 350 (220–480) | 1000 (600–1400) |
| Max daily mortality | 8 (5–12) | 24 (15–33) |
| Peak time-window | 15 May – 3 June | 1 June – 20 June |

**Figure 4.**
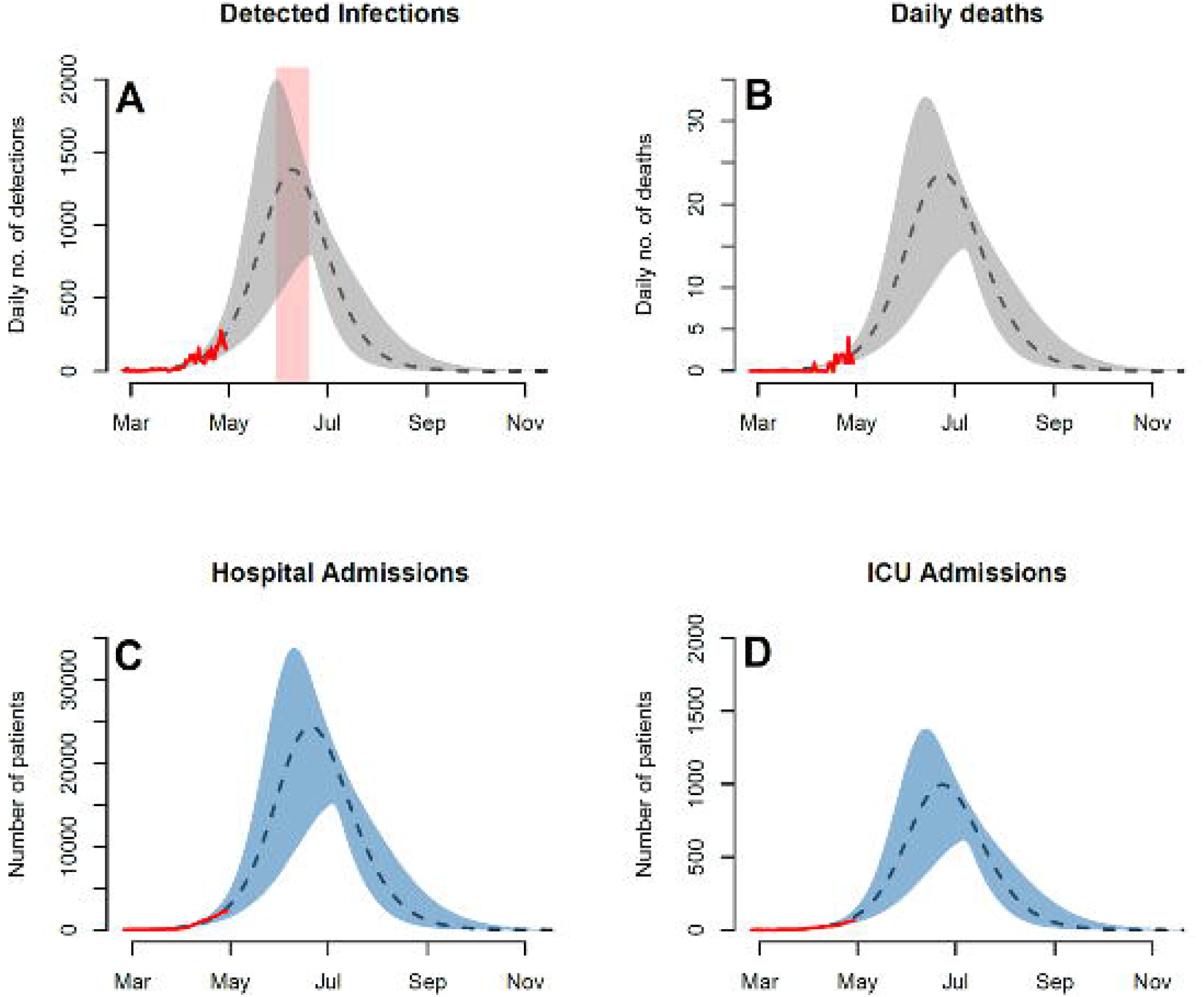
Tripling the size of the unprotected susceptible population. Peaks of forecasted trajectories are approximately tripled in size. The projected time-window of the peak is delayed by 2-weeks and widened (3-week period). Observed and projected daily numbers of A) incident infections, B) death cases, C) general hospital admissions and D) ICU admissions. Red rectangular ribbon highlights the projected time-window of the epidemic peak. Red lines represent the reported data. Black dashed lines represent model projections based on MLE of unknown parameters with shaded ribbons representing 95% credible interval on new observations. We note here that the observed cases and their projections only represent a fraction of the actual and model prevalence. This is based on our assumption of under-reporting and the presence of asymptomatic individuals in the

## DISCUSSION

We developed a mathematical modeling framework for real-time tracking and forecasting of the epidemic outbreak of COVID19 in Kuwait and the associated burden on the healthcare system. This quantitative framework is further employed to evaluate how the control measures implemented in Kuwait may have influenced the epidemic burden. Such tools can derive important policymaking decisions. Insight from Kuwait can potentially inform similar policies, programs and public health measures taken by other developing countries.

Our results suggest that the early gradual and stringent control measures in Kuwait had the effect of delaying and lowering the intensity of the outbreak by protecting a large fraction of the population. Despite the fact that the country has been under lockdown since March 22^nd^, 2020, our model indicates that the effective reproduction number (R_t_) remained unchanged. In principle, such control measures are implemented to achieve a sufficient reduction in the effective reproduction number during an outbreak. This may be explained by the reported outbreaks amongst migrant workers. Kuwait has a considerably heterogenous population with 60% comprised of non-nationals. A large proportion of these workers cluster in certain areas and live in cramped dormitories with poor and unsanitary housing conditions. Strict social distancing measures may not be implementable nor have the same effect on these subpopulations. Hence, this could exacerbate the transmission of the infection in the presence of lockdowns. It is therefore imperative, for the epidemiological understanding of the disease distribution, to perform demographic studies that aim to extract contact structure matrices and establish how different control measures may or may not affect heterogenous transmission rates [23]. This is not only applicable to the State of Kuwait, but also the Arabian Gulf states, Singapore and other countries with somewhat similar demographic profiles.

Additionally, our results indicate that the COVID-19 outbreak in Kuwait is on course to accelerate further in the next few weeks, which is consistent with the recent trends associated with expanded testing and contact tracing. Indeed, our model analysis of the projected epidemic trend indicates that hospitals may need to prepare for admitting around 12000 patients of which 500 may need critical care.

This work has a number of limitations. First, we did not have detailed data on age structure, nor the risk associated with comorbidities. Younger individuals have lower risk of mortality and morbidity from COVID-19, while older people with comorbidities will have significantly higher case fatality ratio. If most of the recorded cases in Kuwait were young, then this might have led us to overestimate the results. In contrast, if the older or comorbid population was higher in Kuwait, we would be underestimating the results. The direction of the bias is critical when adopting this model. Secondly, many parameters were derived from unpublished data obtained from local hospitals in Kuwait. Although the parameters may change as the data is growing, we do not anticipate significant departure from the values presented in this work. Thirdly, we did not have demographic information. Careful consideration should be taken with regards to population heterogeneity, especially migrant workers. Further investigation with geospatial mapping to understand epidemic clusters is warranted. Finally, the complemented R code and real data from Kuwait should not be uncritically applied without careful tailoring to specific study settings and revisiting of the assumptions.

## Conclusions

COVID-19 poses significant public health challenges to many developing countries including Kuwait. We have shown that stringent control measures can effectively delay and lower the intensity of the outbreak. However, they might not be sufficient to completely halt the transmission of the disease in the presence of certain structural restrictions pertaining to population and demographic factors. In turn, this highlights an urgent need for a systematic reassessment of public health interventions to account for demographic heterogeneities. Such an assessment needs to be supported by a modeling tools to monitor the impact on the outbreak progression. In particular, our model can serve as a public health tool for decision makers to guide in the control of the current outbreak, even in the absence of critical population health data. This replicable tool can also be used to anticipate effective future measures should a second wave re-emerge in Kuwait and other developing countries. In addition, it can serve as a public health tool to track and control the current outbreak.

## Data Availability

The data that support the findings of this study are openly available in [An interactive web-based dashboard to track COVID-19 in real time] at [https://www.thelancet.com/journals/laninf/article/PIIS1473-3099(20)30120-1/fulltext], reference number [3].

## Acknowledgments

The authors would like to thank Fatima Khadada from the Department of Medicine (University of Toronto) for helpful comments on the manuscript, and Saud Alzaid from the Department of Surgery (Kuwait University) for valuable input in the formulation of the ICU dynamics of the model.

## Authors Contribution

AA was involved in the conception and design of the study, analysis and interpretation of data and drafting the article. HA was involved in acquisition, analysis and interpretation of data and drafting the article. BA was involved in analysis and interpretation of data. FA1 was involved in interpretation of data, SA was involved in acquisition of data, MJ was involved in acquisition of data, AA2 was involved in acquisition of data, QA was involved in was involved in revising the article critically for important intellectual content and FA2 was involved in analysis and interpretation of data and gave the final approval of the version to be submitted.

## Ethical Statement

Ethical approval was not required for this work.

